# The SARS-CoV-2 B.1.351/V2 variant is outgrowing the B.1.1.7/V1 variant in French regions in April 2021

**DOI:** 10.1101/2021.05.12.21257130

**Authors:** Bénédicte Roquebert, Sabine Trombert-Paolantoni, Stéphanie Haim-Boukobza, Emmanuel Lecorche, Laura Verdurme, Vincent Foulongne, Mircea T. Sofonea, Samuel Alizon

## Abstract

SARS-CoV-2 variants threaten our ability to control COVID-19 epidemics. We analyzed 36,590 variant-specific RT-PCR tests performed on samples collected between April 12 and May 7, 2021 in France to compare variant spread. Contrarily to January to March 2021, we found that the V2 variant had a significant transmission advantage over V1 in some regions (15.1 to 16.1% in Île-de-France and 16.1 to 18.8% in Hauts-de-France). This shift in transmission advantage is consistent with the immune evasion abilities of V2 and the high levels of immunization in these regions.

---

‘Variants of concern’ (VOC) are SARS-CoV-2 phenotypically distinct lineages that are associated with major epidemiological and/or clinical shifts. Variant V1 from lineage B.1.1.7 is currently causing the majority of infections in Europe and North America [1], whereas variant V2 from lineage B.1.351 is dominant in South-Africa [2], and variant V3 from lineage P.1 dominates in Brazil and South America [3]. The outcome of the (indirect) competition between variants is an open question. In France, the early introduction of the V2 variant in some regions makes it particularly important to monitor the spread of different variants [4]. Since January 2021, the national guideline is to test all positive samples with an additional RT-PCR to detect implicated variants [5,6]. Since April 2021, the guideline for variant-specific RT-PCRs is to target the N501Y and E484K mutations.

## Data

We use the ID^™^ SARS-CoV-2/N501Y/E484K Quadruplex assay from ID Solution to test N=53,687 SARS-CoV-2 positive samples collected between Monday April 12 and Friday May 7 2021 in 13 French regions, mainly from the Ile-de-France region (47.9%). Some of the samples (7.95%) originate from hospitals (mostly hospitalized patients). We only analyzed data from individuals aged from 5 to 80 years old to minimize the weight of preschool children and aged care facilities in the analysis. 17.3% of the tests could not be interpreted, because of Ct value too high to keep an equal sensitivity for the N501Y and E484K targets, and were removed from the analysis. Therefore, to avoid biasing the variant screening, all tests with cycle thresholds (Ct) values above 30 were ignored (28%). Overall, we analyze 36,590 tests, i.e. 68.2% of the all the tests performed (Table 1). Raw proportions are shown in Figure 1.

**Table 1.** Characteristics of the N=36,590 samples analyzed via variant-specific RT-PCR. The interquartile range (IQR) shown is the 95% confidence interval.

| **variable** | **value** | **N (%) or IQR** |
| --- | --- | --- |
| age | --- | 38 [10,73] |
| sampling location | *general population (ref.)* | 34,022 (93%) |
|  | *hospital* | 2,568 (7%) |
| variant | *N501Y- & E484K-* | 662 |
|  | *N501Y+ & E484K-* | 31929 |
|  | *N501Y- & E484K+* | 647 |
|  | *N501Y+ & E484K+* | 3352 |
| Ct | --- | 22.1 [14.9,29.4] |
| sampling date | --- | Apr 21 [Apr 12,May 6] |
| region | *Normandie (ref.)* | 6288 |
|  | *Centre-Val de Loire* | 1962 |
|  | *Hauts-de-France* | 4684 |
|  | *Ile-de-France* | 16922 |
|  | *Occitanie* | 409 |
|  | *Provence-Alpes-Côte d'Azur* | 3789 |
|  | *Bourgogne-Franche-Comté* | 454 |
|  | *Nouvelle-Aquitaine* | 1444 |
|  | *other* | 639 |

**Figure 1:**
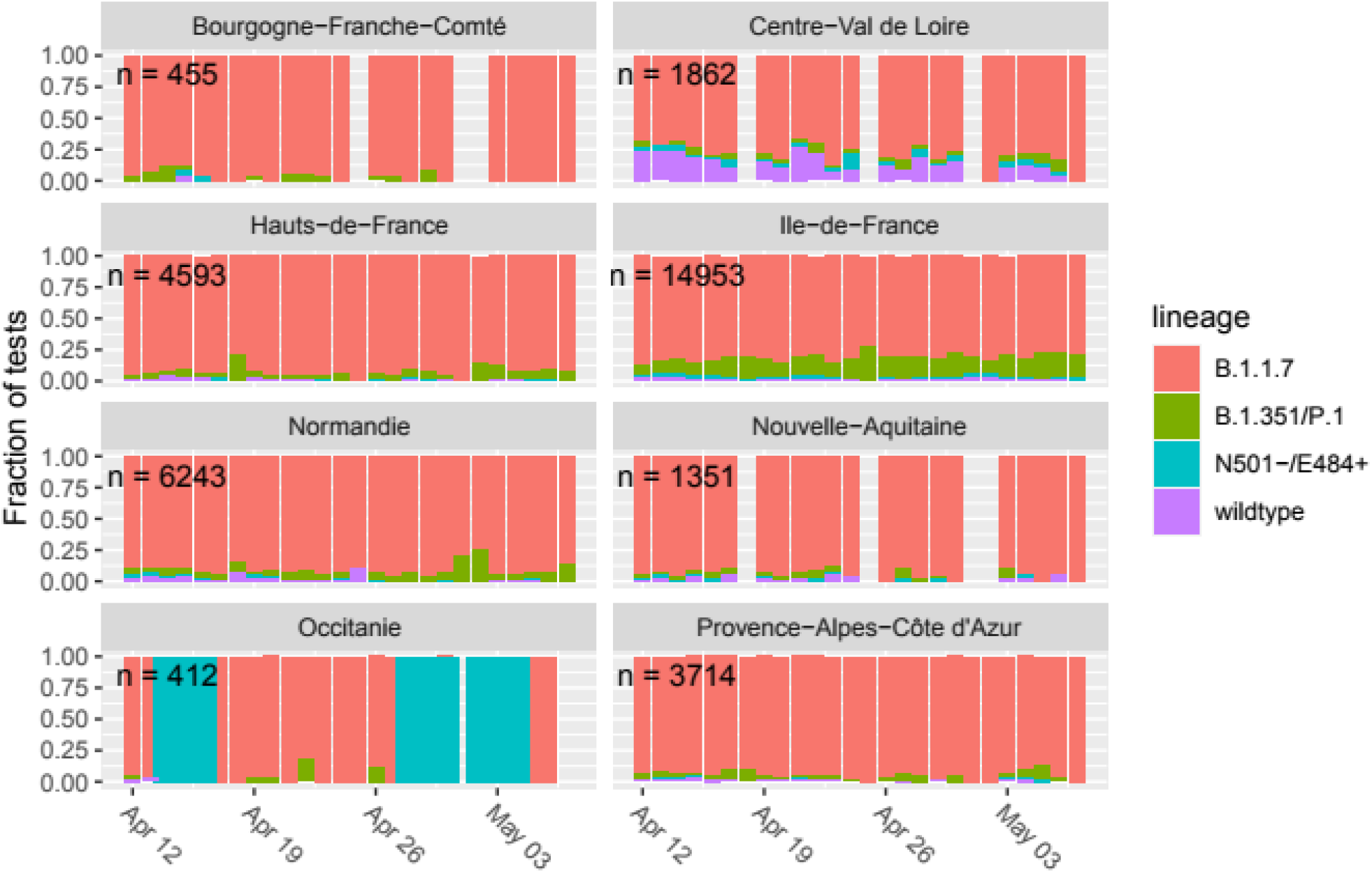
Raw daily frequencies of variant-specific test results in French regions. The number of tests performed is indicated in each panel. Regions with few tests can exhibit strong variations in frequencies. Wild type corresponds to N501Y-/E484K-, B.1.351/V2 to N501Y+/E484K+, B.1.525 to N501Y-/E484K+, and B.1.1.7 to N501Y+/E484-. See supplementary figure S1 for the raw numbers instead of the proportion.

### Sequencing profiles

The specificity of the variant-specific RT-PCR we used is limited since they only target 2 mutations. To gain additional insights, we sequenced all the samples collected on the same day (March 30) for which the Ct was lower than 28 using Twist Libraries and Illumina sequencing (currently being uploaded to GISAID).

These 478 samples showed a majority of viruses from the B.1.1.7 / V1, lineage (79.1%). The other prevalent lineages were B.1.351 / V2 (7.9%), B.1.525 (4.4%), and B.1.214 (2.3%). Other lineages represented less than 2% of the samples (lineage P.1 / V3 being below 1%). Therefore, in the following, samples with only the N501Y mutation are treated as V1, samples with both mutations as V2, samples with only the E484K mutation as B.1.525, and samples with no mutation as wild types (B.1.214 being rare).

### Lineage spreading in France

We used a multinomial log-linear model (the `multinom` function from the `nnet` R package) to analyze the lineage variable (V1 being the variant of reference). The explanatory variables were the patient age, individual location (in a hospital or not), and the interaction between the sampling region and date.

The multinomial model revealed differences between lineages (Table 1). In terms of age, we find that older patients tend to be more infected by V1 (our reference), than by V2 or B.1.525. In hospital settings, we find an over-representation of V2 compared to V1. When analyzing region-specific temporal trends, we found that the odds of being infected by a wild type or a B.1.525 virus were either identical or lower than the one of being infected by V1. Conversely, in Île-de-France, and to a lesser extent in Hauts-de-France and Nouvelle-Aquitaine, we find that the risk of being infected by V2 instead of V1 significantly increases with time.

**Table 1.** Odds ratios regarding the risk of lineage detection estimated using a multinomial log-linear model. The reference lineage in the analysis is B.1.1.7 (N501Y+/E484K-), i.e. the variant of concern V1. Only factor values with a least one significant effect are shown. P-values are obtained using a 2-tailed z test (symbols indicate p-values lower than 10^−3^ for ^***^, than 10^−2^ for ^**^, and than 0.05 for ^*^).

| **Factor** | **Value** | **wild type**<br>(N501Y-/E484K-) | **B.1.351 / V2**<br>(N501Y+/E484K+) | **B.1.525**<br>(N501Y-/E484K+) |
| --- | --- | --- | --- | --- |
| age (per year) |  | n.s. | 0.95** | 0.86** |
| location | *Non-hospital (ref.)* | --- | --- | --- |
|  | *hospital* | n.s. | **1.56\*\*\*** | n.s. |
| date x region | *Normandie* | 0.54*** | n.s. | 0.69*** |
|  | *Centre-Val de Loire* | 0.26*** | n.s. | n.s. |
|  | *Hauts-de-France* | 0.72** | **1.14\*** | n.s. |
|  | *Ile-de-France* | 0.74*** | **1.42\*\*\*** | 0.81*** |
|  | *Nouvelle-Aquitaine* | n.s. | **1.25\*** | n.s. |
|  | *Occitanie* | n.s. | n.s. | n.s. |
|  | *Provence-Alpes-Côte d'Azur* | n.s. | n.s. | n.s. |
|  | *other* | n.s. | n.s. | n.s. |

### Transmission advantage

Trends from the multinomial model should be treated with caution because of auto-correlation issues. Therefore, to investigate the temporal trends, we used the method described in [5] and fitted a logistic growth model to the fitted values of a generalized linear model (GLM) with 4 factors (sampling date, sampling department, and individual age) on the data sampled outside hospitals. For simplicity, we tested the transmission advantage of V2 compared to V1 and neglected the other strains in the analysis. We performed the analysis only in the Île-de-France, Hauts-de-France, and Nouvelle-Aquitaine regions. In the absence of adequate data to infer a serial interval in France, we use the one from [7].

We found a transmission advantage of 15.8% (95% confidence interval: [15.1,16.1]%) in Île-de-France and 17.1% (95%CI: [16.1,18.8]%) in Hauts-de-France (Figure 2). In Nouvelle-Aquitaine, the logistic growth was not significant.

**Figure 2:**
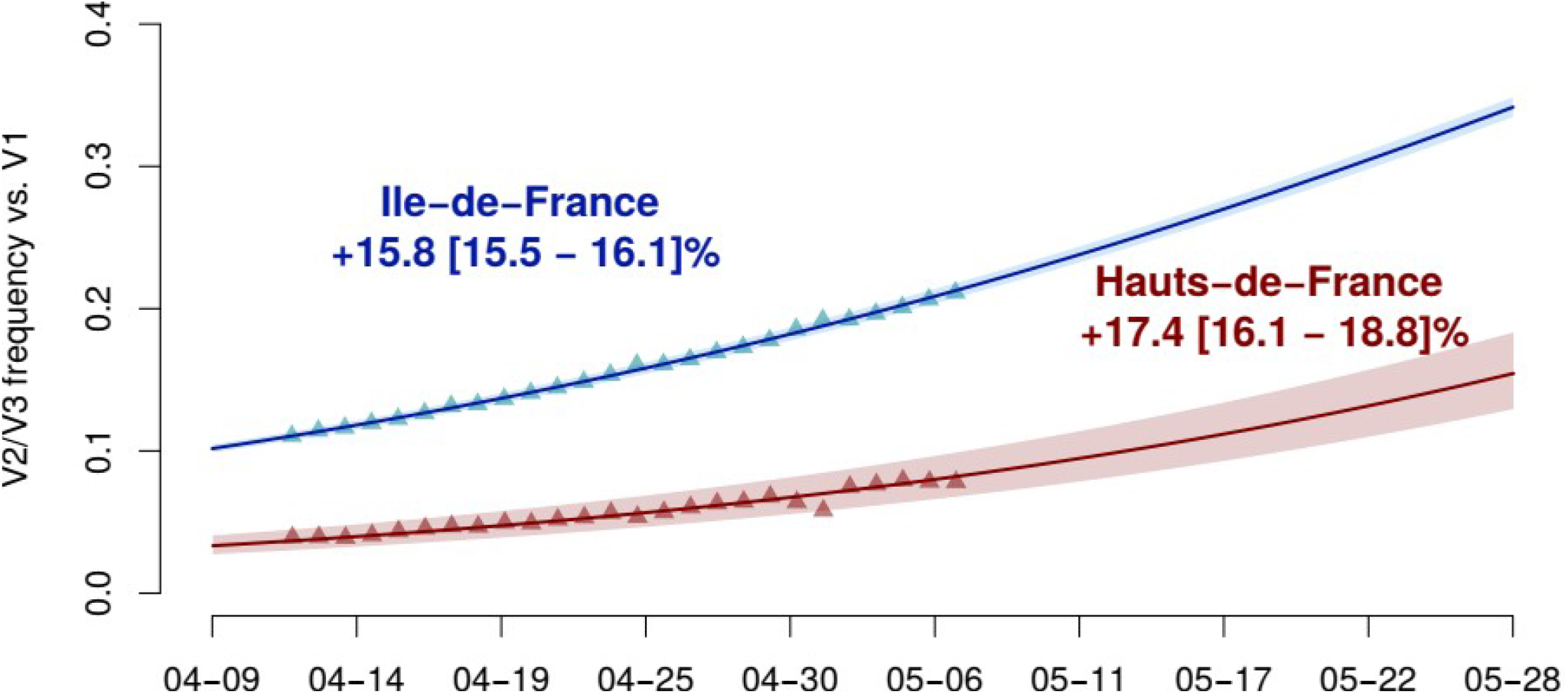
Estimated proportion of V2 with respect to V1 (other lineages are excluded) in Île-de-France (blue) and Hauts-de-France (red). The triangles indicate the GLM-fitted values and the line is the output of the logistic growth model estimation. The top figures indicate the estimated transmission advantage of V2 with respect to V1 and the 95%-confidence interval.

## Discussion

When analyzing variant-specific testing spanning from January to March 2021, we found that V1’s transmission advantage to the wild type was larger than that of V2 [8]. During April 2021, in at least two French regions, this trend appears to have shifted with V2 spreading more rapidly than V1. The V2 variant has known immune evasion properties [9,10]. Therefore, Île-de-France being one of the French regions the most impacted to date by the epidemic, it is sensible that we detect the shift in transmission advantage there because the proportion of individuals with natural immunity being high. Vaccination can also be expected to favor immune escape mutants [11] but the coverage is more homogeneous among French regions. These results call for more detailed analyses regarding the link between the transmission advantage of the V2 variant and the proportion of the population immunized in different French regions. Finally, note that France has entered a third national lock-down on April 3^rd^, which means that most of the tests analyzed here have been performed in a declining epidemic [12]. However, it is unlikely that the lock-down *per se* would affect differently the transmission of the variants. Given the high COVID-19 incidence in France and the uncertainties regarding the lock-down lifting [12], these results call for particular care to avoid another epidemic wave in the country.

## Data Availability

Data will be made available upon manuscript publication.

## Ethics

This study has been approved by the Institutional Review Board of the CHU of Montpellier and is registered at ClinicalTrials.gov with identifier NCT04738331.

## Acknowledgements

We thank the ETE modelling team for discussion, as well as the CNRS, the IRD, the ANR, and the Région Occitanie for funding (PHYEPI grant). We thank the Infectious diseases department in Cerba laboratory, technical and IT teams for their technical assistance.

---

**Figure S1:**
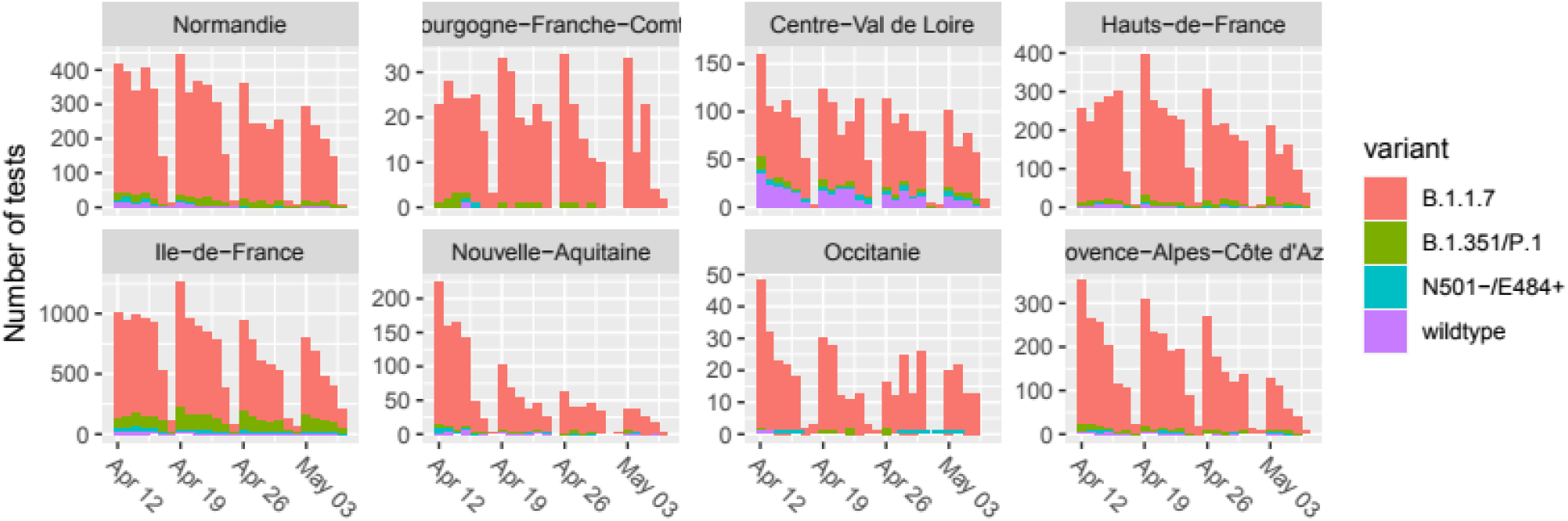
Raw daily occurrences of variant-specific test results in French regions. The number of tests performed is indicated in each panel. Wild type corresponds to N501Y-/E484K-, B.1.351/V2 to N501Y+/E484K+, B.1.525 to N501Y-/E484K+, and B.1.1.7 to N501Y+/E484-.

